# Psychiatric comorbidities in substance use disorders: Sex-based differences in a national real-world clinic sample

**DOI:** 10.1101/2025.09.11.25335563

**Authors:** Eduardo R. Butelman, Yuefeng Huang, Rita Z. Goldstein, Nelly Alia-Klein

**Author notes:** **Corresponding Author:**, Eduardo Butelman PhD, Neuropsychoimaging of Addictions and Related Conditions, Department of Psychiatry, Icahn School of Medicine at Mount Sinai, New York NY 10029. **Disclosures:** The authors have not disclosures related to this study. **Funding:** This study was funded from grants from the National Institute on Drug Abuse, which the authors gratefully acknowledge.

## Abstract

**Objective:** Substance use disorders (SUD) are associated with risk of psychiatric comorbidities, with inconsistent sex differences, across studies. The objective of this study is to determine odds of psychiatric comorbidities based on sex, in persons with either primary opioid, alcohol or cannabis dependence diagnoses, from a national clinical data set.

**Methods:** This is a cross-sectional study of data from state-funded and state-run mental health programs in 2022 (Mental Health Client-Level Data, from the US Substance Abuse and Mental Health Administration). Data was obtained from adults (age ≥18) with a primary diagnosis of either opioid (n=28,808), alcohol (n=23,281) or cannabis dependence (n=5,961). Individuals with each SUD were examined for psychiatric comorbidity outcomes, based on secondary diagnoses of either anxiety, bipolar, depression, schizophrenia or other psychotic disorders (SPD), or trauma and stressor-related disorders, versus no comorbidity. Data were analyzed with multinomial logistic regressions, examining sex, race, ethnicity and age as predictors.

**Results:** Males with primary diagnoses of either opioid, alcohol or cannabis dependence had lower adjusted odds of anxiety, bipolar, depression and trauma/stressor disorders, compared to females. However, males with opioid or cannabis dependence had higher adjusted odds of SPD, compared to females. Adjusted analyses also detected associations of race and ethnicity with specific comorbidities.

**Conclusions:** In this recent national clinical data set, there are specific sex-based differences in specific psychiatric comorbidities for each of these three SUDs. Future studies should examine biopsychosocial mechanisms that underlie these differences, with the goal of improving personalized care.

## Introduction

Psychiatric comorbidities are frequent in individuals with substance use disorders (SUD), including opioid, alcohol or cannabis use disorders(1–3), and can affect clinical trajectories and treatment outcomes(3, 4). Recent estimates indicate that only 7% of adults with such dual diagnoses receive integrated treatment, and optimization of personalized care and clinical resources therefore remains a critical need(5). Several cohort studies and surveys have reported that some psychiatric comorbidities (including anxiety and trauma/stress disorders) are more common in females than males(6, 7), whereas others (e.g., schizophrenia) may show the opposite profile(7). The potential causes for such sex disparities are unclear, and may be based on biological(8–10), behavioral or social (11) mechanisms. Race and ethnicity(12, 13), potentially operating through several underlying mechanisms(14, 15), are also associated with disparities in SUD trajectory, treatment and outcomes, based on cohort studies and survey data(16–19). It is therefore important to study race and ethnicity - based heterogeneities, as part of a broad study of sex-based psychiatric comorbidities in large and diverse clinical populations.

The underlying mechanism(s) for co-occurrence of opioid, alcohol (ethanol) and cannabis use disorders and psychiatric comorbidities are areas of active study(3, 4, 20, 21). Furthermore, while mu-opioid receptor (MOR) agonists such as fentanyl or heroin, ethanol and cannabinoids act directly on different receptor targets, they can have shared downstream neurobiological effects(22–24), which may underlie conditions such as anxiety, mood, schizophrenia, and trauma/stress disorders(25, 26), including instances of sexual dimorphism(10, 27). More broadly, formal research studies and surveys alone may not fully capture sex-based heterogeneities in comorbidities, due to specific inclusion/exclusion criteria, relatively small sample size of persons with each SUD, differential representation based on race or ethnicity, and self-reporting of stigmatized conditions(12, 28–31). Therefore, large-scale real-world clinical data sets(32, 33) are an especially valuable resource(12, 34). Based on this premise, the primary goal of this study was to analyze sex-based heterogeneities of major psychiatric comorbidities in individuals with primary opioid, alcohol or cannabis use disorders, in a large and recent national clinic sample from the United States.

## Methods

### Study Design

Cross-sectional study, examining sex differences in the prevalence of psychiatric comorbidities as outcomes in persons with specific SUD as their primary mental health diagnoses, adjusting for race, ethnicity and age. **Inclusion Criteria:** Individuals with a primary mental health diagnosis of “alcohol or substance use disorders” (variable MH1) who were at least 18 years of age. **Exposure:** Individuals who had one of the substance use diagnosis (variable SUB) of either: opioid dependence, alcohol dependence or cannabis dependence (the provided DSM-IV nomenclature); the remainder were excluded. Other SUD such as cocaine or methamphetamine dependence were not studied, due to relatively low “n” in the data set. Data were further filtered as described below (see “Variable Selection Strategy”). **Outcome:** The outcome was presence of a psychiatric comorbidity (variable MH2, secondary mental health diagnosis) of either anxiety, bipolar, depressive (termed “depression” hereafter), schizophrenia or other psychotic disorders (SPD), or trauma and stressor-related (termed “trauma” hereafter) disorders, versus absence of any psychiatric comorbidity. The aforementioned comorbidity categories were the most prevalent in this data set. Individuals with other comorbidities were excluded from the main analyses, due to their heterogeneity and relatively lower prevalence (see also Participant Selection Table in the Supplement).

### Data Source

Data are from the 2022 Mental Health Client-Level Data (MH-CLD) from the United States Substance Abuse and Mental Health Services Administration (SAMHSA), which contains publicly available, observational de-identified data, and are not considered Human Subjects Research, as determined by the Institutional Review Board of the Icahn School of Medicine at Mount Sinai. Data are obtained from state-funded or state-run mental health facilities in the United States (excluding American Samoa, Federated States of Micronesia, Guam, Marshall Islands, Virgin Islands and Maine). Raw data, full methodological features, description of confidentiality and privacy protections, variable descriptions and frequencies are publicly available: https://www.samhsa.gov/data/.

### Variable selection strategy

The variable selection and filtering strategy, and participants at each stage are summarized in the Participant Table (Supplement). From the individuals who had either opioid, alcohol or cannabis dependence primary diagnoses, we selected those who had a secondary mental health diagnosis (variable MH2) of either anxiety, bipolar, depressive, schizophrenia or other psychotic disorders (SPD), or trauma and stressor-related disorders (thus including post-traumatic stress disorder). As a comparison (reference) group without psychiatric comorbidity, we filtered individuals who only had the “alcohol or substance use disorders” primary diagnosis as above, and no other diagnosis (using variable NUMMHS, which reports the number of valid mental health diagnoses). Variables for living arrangements and employment were not entered in the analysis, since these data do not specify whether information was obtained at the end of the reporting period (for continuing clients) or at discharge, thus introducing a potential confounder(32). **Covariates:** Sex (Male or Female), race (three categories: African-American / Black, American Indian / Alaska Native, or White), ethnicity (re-coded as a binary variable: hispanic/latino or non-hispanic/latino), and age (re-coded in consecutive age bins). Individuals of other racial categories were not analyzed due to low prevalence(35). Persons were excluded if they were not in the main U.S. Census regions, due to low prevalence. **Missing data:** Among the variables under study, sex and age had missingness rates <1%; ethnicity and race had missingness of ≈20%. Case-wise deletion was therefore used when there were missing values in these variables. The education variable had a greater rate of missingness (76%) and was therefore not used in the main analyses; imputation was not attempted.

### Statistical analyses

The p=0.05 level was selected as alpha for the study. Statistical analyses were carried out with GraphPad Prism or TIBCO Statistica Data Science Workbench. **Univariate analyses** were used to examine demographics with Chi^2^ contingency analyses. Chi^2^ contingency analyses were also used to examine proportions of persons with a specific comorbidity (e.g., by sex, race or ethnicity); 95%CI of the proportions were calculated with the Wilson-Brown method(36). **Multivariate analyses:** The primary outcomes were five mutually exclusive psychiatric comorbidity diagnostic groups (i.e., presence of anxiety, bipolar, depression, SPD and trauma/stressor) versus a reference group without a psychiatric comorbidity. For this multi-class classification design, these analyses were carried out with three multinomial logistic regressions (i.e., examining individuals with opioid, alcohol or cannabis dependence separately), adjusting for the covariates above. Beta-parameters, adjusted odds ratios and 95%CI are reported, as well as likelihood ratio test (type 3), examining unique contributions of each predictor.

### Follow-ups and sensitivity analyses

1) Psychiatric comorbidities (variable MH2) other than those mentioned above had lower prevalence. In a sensitivity analysis, these other comorbidities combined were examined as a further composite outcome category. 2) A follow-up subset analysis including the education variable as a covariate was also carried out; because of the relatively high rate of missingness for this variable, this analysis was carried out only in the largest SUD groups (opioid and alcohol dependence).

## Results

### Demographics

Table S1 shows the distribution of demographic variables and outcomes in each of the three SUD groups. The relative frequency of males was greater than that of females, in each SUD group. Persons in the white racial category were the most frequent in each of the SUD groups. More broadly, the rank order of frequencies for persons in the African-American/Black (AA) and American Indian/Alaska Native (AI) racial categories differed across the SUD. Likewise, the relative distribution of persons of hispanic ethnicity differed across the SUD. Age distribution was not homogeneous in the SUD groups. Thus, in individuals with opioid dependence and cannabis dependence, the most prevalent age bin was 25-34, and declining at older age bins. However, for individuals with alcohol dependence, the most prevalent age bin was older (age ≥55). **Overall distribution of psychiatric comorbidities in individuals with different SUD:** Full data on the relative frequency of comorbidities in all individuals (males and females combined) are in Tables S2-4, and summarized in Figure S1. For individuals with opioid dependence, the two most prevalent comorbidities were anxiety and depression (14.7% and 13.5%, respectively), and the least prevalent was SPD (2.0%). This general profile was also observed in individuals with alcohol dependence, in whom the two most prevalent comorbidities were again anxiety and depression (12.0%, 21.8%, respectively) and and the least prevalent was SPD (5.4%). However, for individuals with cannabis dependence, the two most prevalent comorbidities were depression and SPD (18% and 13.4%, respectively), and the least prevalent was trauma disorders (8.5%).

### Psychiatric comorbidities in persons with different SUD - univariate analysis

We visualized the univariate distribution of psychiatric comorbidities stratified by sex in each SUD, as shown in Fig. 1A-C (based on Tables S2-4). Omnibus Chi-square tests for psychiatric comorbidity status in males and females were significant for each of the three SUD (Tables S2-4). We also visualized univariate comorbidity data stratified by racial group (Fig. 1D-F) and ethnicity SUD (Fig 1G-I). Omnibus Chi-square tests for psychiatric comorbidity status based on race and ethnicity were also significant for each of the three SUD (Tables S2-4).

**Figure 1.**
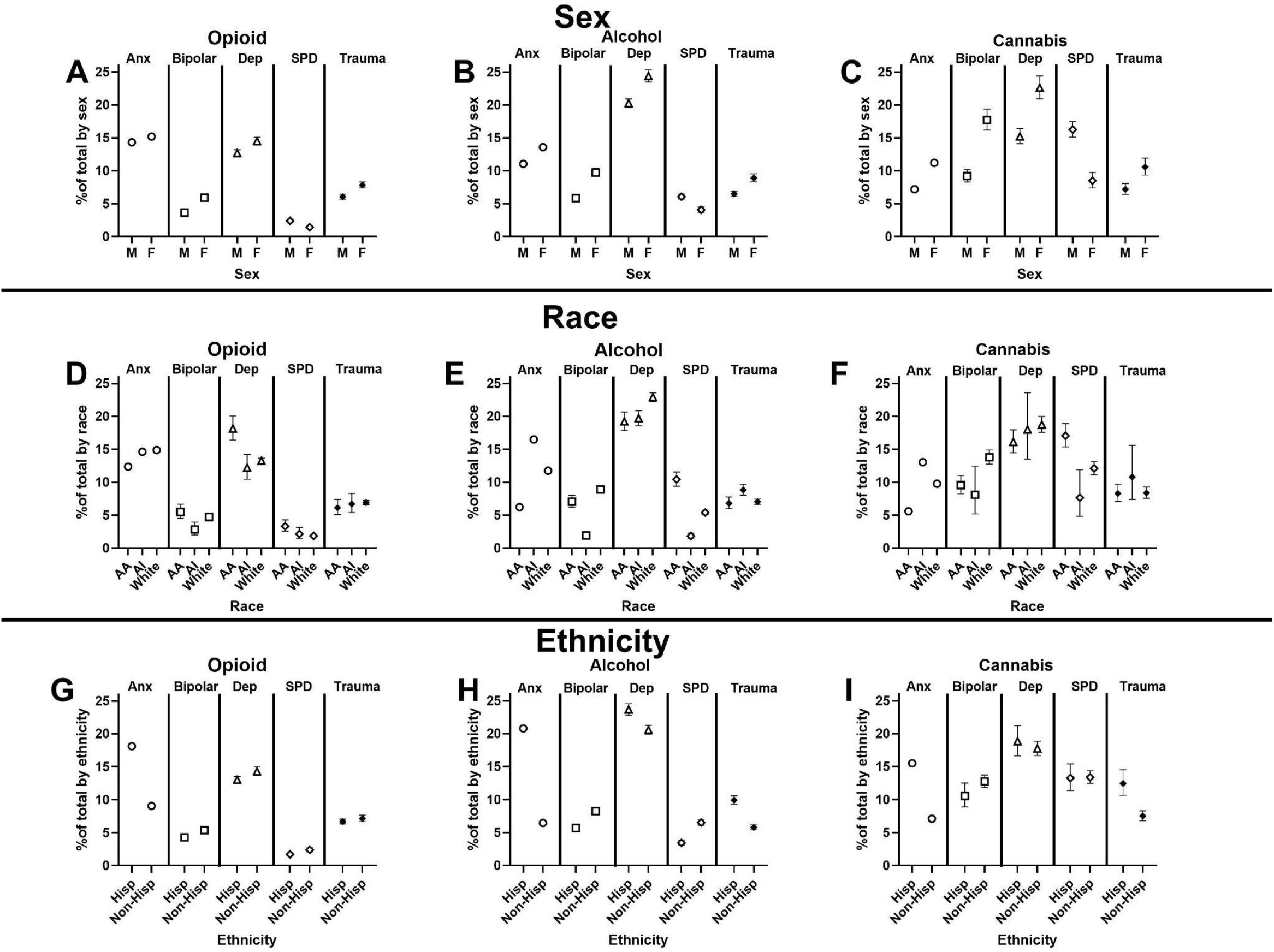
Psychiatric comorbidities based on sex (A-C), race (D-F) and ethnicity (G-I), among individuals with opioid, alcohol or cannabis dependence. Data show the percent of individuals with each primary SUD diagnosis who have a specific psychiatric comorbidity; 95%CI of the proportions were calculated with the Wilson-Brown method(36) (some 95%CI bars are not visible, as they fall within the symbols). Percent data are based on the total sample, including those without a comorbidity (full data in Tables S2-4, including significant omnibus Chi-square tests for each panel). Abbreviations: Anx: Anxiety; Dep: Depression; SPD: Schizophrenia or other psychotic disorders; AA: African-American or Black; AI: American Indian or Alaska Native; Hisp: Hispanic; Non-Hisp: Non-Hispanic. These data were then analyzed in multinomial logistic regressions (Figs 2-4, and Tables S5-7).

### Multivariate analyses

Data were then analyzed in multinomial logistic regressions, separately for each SUD. The presence of the different comorbidities was used as the outcome, versus no comorbidity. Sex, race, ethnicity and age were examined as predictors; full regression parameters are in Supplementary tables S5-7. Results are summarized below.

### Opioid Dependence

Among individuals with opioid dependence (Fig. 2 and Supplement Table S5), males had smaller adjusted odds of anxiety, bipolar, depression, and trauma disorder outcomes, compared to females. However, males had greater adjusted odds of SPD, compared to females. Also, persons in the AA race category had greater odds of bipolar depression and SPD, compared to the reference category (white). Persons in the AI race category had smaller odds of bipolar disorders and depression, compared to the reference category (white). Persons of hispanic ethnicity had greater adjusted odds of anxiety, compared to the reference category (non-hispanic/latino), but smaller adjusted odds of bipolar and SPD. Odds of anxiety, bipolar, depression and trauma disorders were greatest in the youngest age category (18-24) versus the oldest age category (≥55), used for reference.

**Fig. 2.**
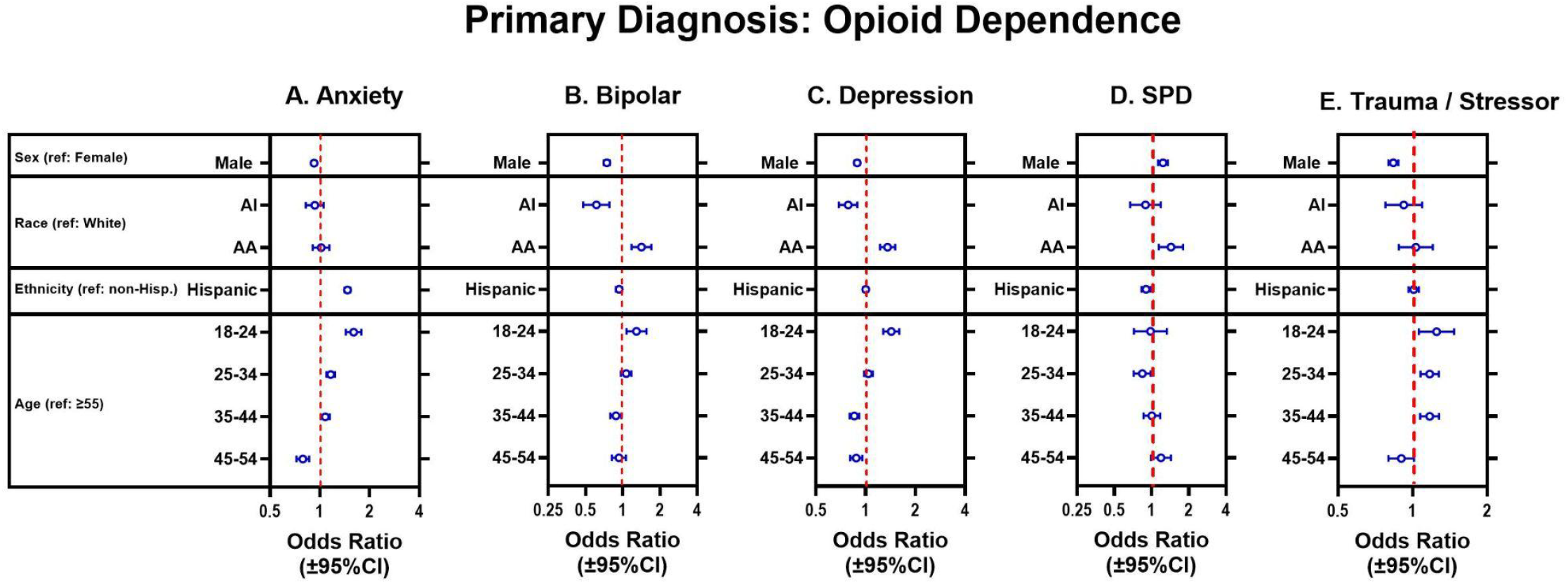
Adjusted odds ratios (and 95%CI) of anxiety, bipolar, depression, schizophrenia or other psychotic disorders (SPD) or trauma/stressor psychiatric comorbidities (panels A-E) in individuals with a primary diagnosis of opioid dependence, from multinomial logistic regression. Note different x-axis scales across panels; full regression parameters are in Supplement Table S5. Ref.: reference category

### Alcohol Dependence

Among individuals with alcohol dependence (Fig. 3 and and Supplement Table S6), males had smaller adjusted odds of anxiety, bipolar, depression, and trauma disorders, compared to females. However, odds of SPD did not differ in males versus females. Persons in the AA race category had greater odds of bipolar, SPD and trauma/stressor disorders, but smaller odds of anxiety, compared to those in the reference category (white). Persons in the AI race category had smaller odds of all the comorbidity outcomes, compared to those in the white category. Persons of hispanic/latino ethnicity had greater odds of anxiety, bipolar, depression and trauma/stressor disorders, compared to the reference category (non-hispanic/latino). There were different age-related profiles in odds of the different comorbidities, compared to the oldest category (age ≥55).

**Fig. 3.**
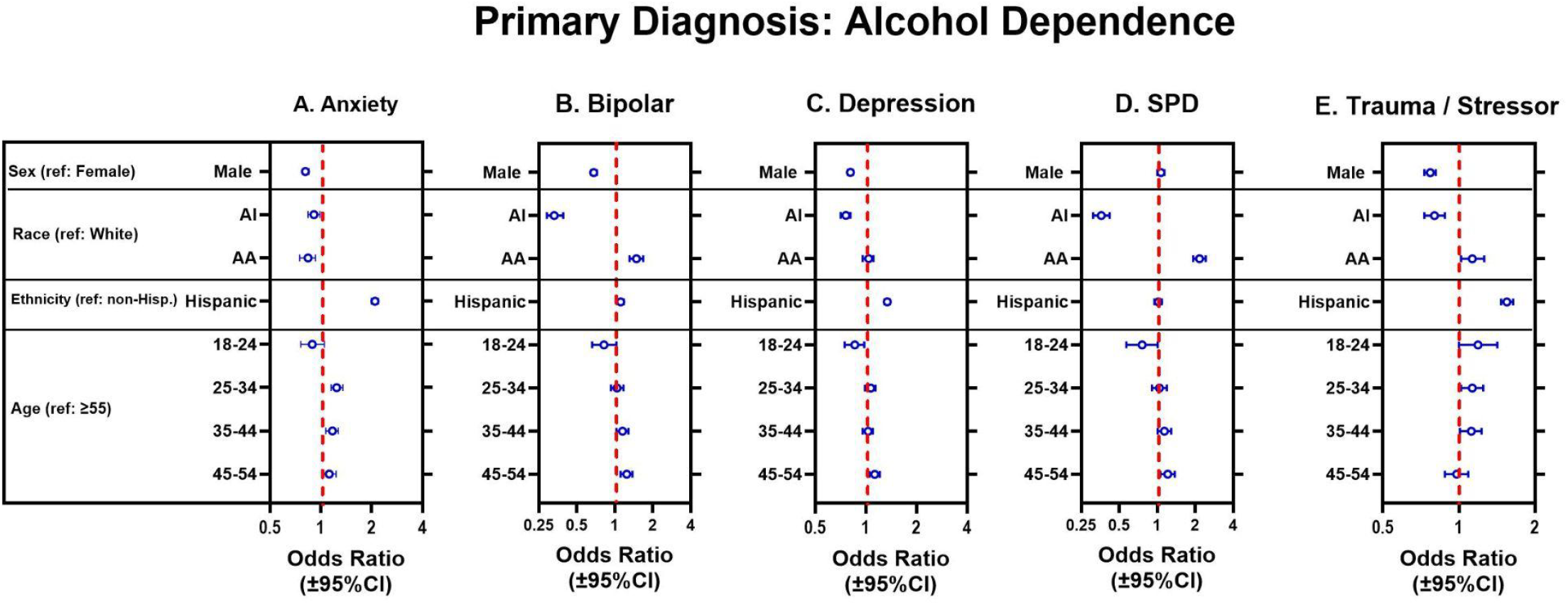
Adjusted odds ratios (and 95%CI) of anxiety, bipolar, depression, SPD or trauma/stressor psychiatric comorbidities (panels A-E) in individuals with a primary diagnosis of alcohol dependence, from multinomial logistic regression. Note different x-axis scales across panels; full regression parameters are in Supplement Table S6.

### Cannabis Dependence

Among individuals with cannabis dependence (Fig. 4 Supplement Table S7), males had smaller adjusted odds of anxiety, bipolar, depression, and trauma/stressor disorders, compared to females. However, odds of SPD were greater in males versus females. Persons in the AA race category had greater odds SPD and trauma/stressor disorders, compared to the reference category (white), but smaller odds of anxiety. Persons in the AI race category (which had a relatively low “n” in this group) had smaller odds of bipolar and SPD compared to those in the reference category. Persons of hispanic/latino ethnicity had greater odds of anxiety, depression, SPD and trauma/stressor disorders, compared to the reference category (non-hispanic/latino). Persons in the two youngest age categories (18-24 and 25-34) had greater odds of anxiety, compared to the oldest category (age ≥55). There were also different odds of depression and SPD in the youngest age category (18-24) versus those in the oldest category (age ≥55).

**Fig. 4.**
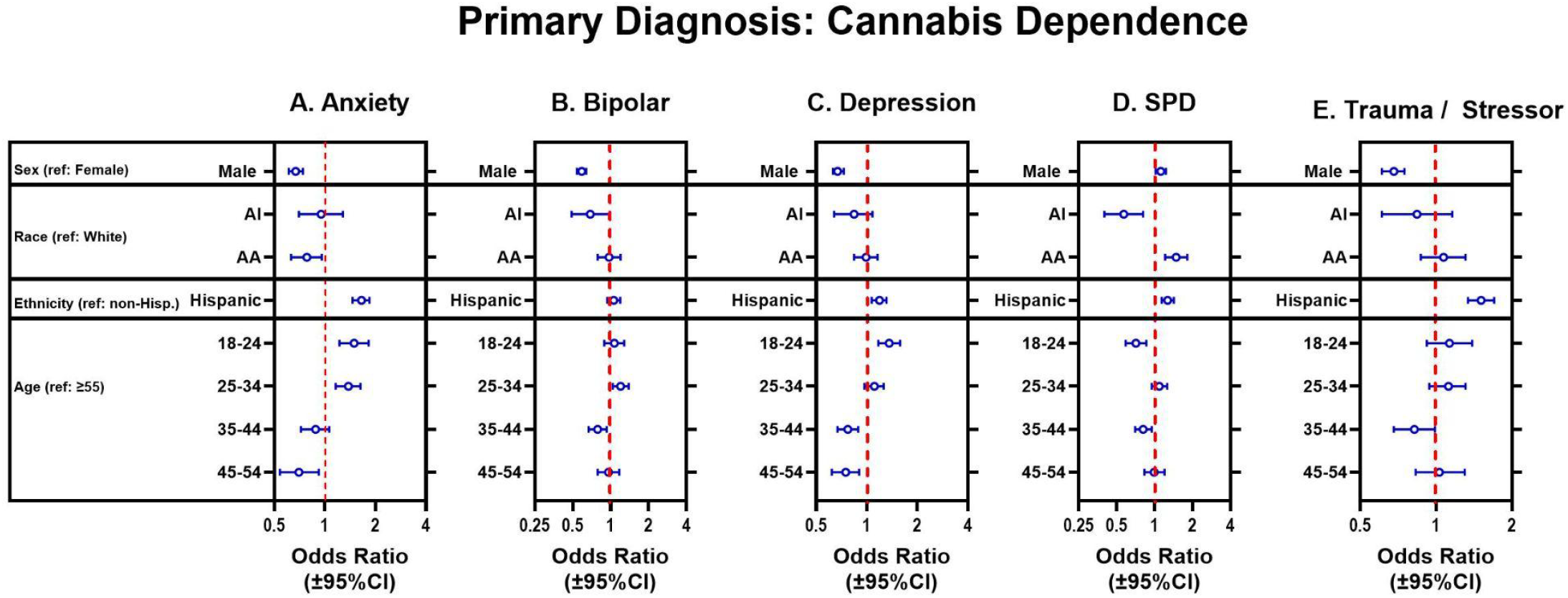
Adjusted odds ratios (and 95%CI) of anxiety, bipolar, depression, SPD or trauma/stressor psychiatric comorbidities (panels A-E) in individuals with a primary diagnosis of cannabis dependence, from multinomial logistic regression. Note different x-axis scales across panels; full regression parameters in Supplement Table S7.

### Follow-ups, sensitivity and subset analyses

**1)** As a follow-up, we examined all other psychiatric comorbidities combined, as a further composite outcome category (each of these other comorbidities alone had relatively low prevalence). This composite category was composed of n=2062, n=2539 and n=649 in opioid, alcohol and cannabis dependence groups, respectively. Addition of this composite comorbidity category to the multinomial logistic models did not result in substantial changes to the findings in Figs 2-4 (not shown). **2)** As another follow-up, we examined whether addition of education as a predictor would affect the findings in adjusted analyses. Due to the relatively high rate of missingness in the education variable, we only examined the two largest SUD groups herein (opioid and alcohol dependence; with data available from n=6,014 and n=8,305, respectively). Education was studied in four categories: 0-8 years of education, 9-11 years, 12 years or equivalent, and >12 years as the reference category. In this follow-up analysis, the sex disparities for anxiety, bipolar, depression and trauma disorders were still significant, after adjustment for education level (not shown). Only the sex disparity that was observed for SPD in the opioid dependence group did not retain significance in this follow-up. Furthermore, most of the disparities observed in persons in the AA racial group survived this adjustment for education, whereas those in the ethnicity variable did not (not shown).

## Discussion

This analysis of a real-world national clinic sample of individuals with primary diagnoses of either opioid, alcohol or cannabis dependence revealed sex differences in adjusted odds of major psychiatric comorbidities. The main finding of this study is a common pattern of *lower* adjusted odds of anxiety, bipolar, depression and trauma comorbidity in males versus females, in each of the three SUD. By contrast, males had *higher* odds for the SPD comorbidity in persons with opioid or cannabis dependence. However, no significant sex difference for SPD was observed in persons with alcohol dependence.

There are several potential biopsychosocial mechanisms that could underlie sex differences in these comorbidities, to be analyzed directly in future studies. These mechanisms could include hormonal and organizational influences on neurobiological functions(37, 38), and also shared downstream mechanisms of the SUD and these comorbidities(39, 40). Furthermore, sex-or gender-based differences in vulnerability to psychological distress, stigma, help-seeking, clinical presentation and available therapeutic resources are also important targets for study(41, 42).

As a secondary focus, adjusted analyses also show that race and ethnicity(12) were also associated with differences in comorbidities in individuals with each SUD. Thus, persons in the AA racial category had higher odds of bipolar, depression, and SPD, compared to the reference category (white). However, persons in the AI racial category with opioid dependence had a qualitatively different pattern of psychiatric comorbidities, with *lower* odds of bipolar disorders, compared to the reference category (white). Persons of hispanic/latino ethnicity with opioid dependence had higher adjusted odds of anxiety, compared to the reference category (non-hispanic/latino), but lower adjusted odds of bipolar disorders and SPD. Overall, these differences based on race and ethnicity may be due to sociocultural features, aspects of psychiatric symptom presentation and diagnosis, or differential trauma/stress exposure(43, 44). As an example, a subset follow-up analysis in opioid and alcohol dependence groups adjusting for education, resulted in decreased significance of odds ratios for anxiety in persons of hispanic ethnicity. Overall, these findings underscore the importance of analyzing race and ethnicity together with sex, in future analyses of psychiatric comorbidity.

### Limitations

**1)** This study contains a relatively large national clinic sample, but may not be representative of individuals who present for care at non-state-run/state-funded facilities(45), or those who do not present for care(5). **2)** Observational studies such as this cannot be used to discern the potential causality links between primary SUD diagnoses and psychiatric comorbidities. **3)** Available data for the comorbidities were in diagnostic groups (e.g, anxiety, bipolar, depression, trauma/stressor disorders, or SPD), and were not further stratified into individual diagnoses (e.g., generalized anxiety disorder, etc). **4)** For simplicity, we only analyzed one comorbidity per person (i.e., the second mental health diagnosis; variable MH2), and not possible further comorbidities. **5)** We only examined persons in the three most prevalent racial categories in the dataset (AA, AI and white)(35). Also, the “n” for persons in the AI racial group was relatively small (especially among those with cannabis dependence), requiring larger follow-ups. **6)** This study did not examine employment or housing variables, because it is unknown if these data were obtained at treatment discharge or at the end of the reporting period, potentially introducing a confounder(32).

### Conclusions and implications

In this national real-world clinic study, we detected sex disparities in specific types of psychiatric comorbidities for opioid, alcohol and cannabis use disorders. These studies expand our previous knowledge from formal studies using different cohorts and methodologies(28, 46), or from survey data. Overall, the present findings underscore the need to understand potential multicausal underpinnings of these sex disparities in the different comorbidities for each SUD(8, 11, 47), with the long-term goal of improving personalized prevention, diagnosis and treatment strategies.

## Supporting information

Results Supplement

Participant filtering table

## Data Availability

Data summaries are presented in supplements. Also all source data are in: https://www.samhsa.gov/data/data-we-collect/mh-cld-mental-health-client-level-data/datafiles

## Abbreviations

95%CI: 95% confidence interval
AA: African-American or black
AI: American Indian or Alaska Native
Anx: Anxiety
AUD: alcohol use disorder
Dep: Depression
Df: degrees of freedom
F: female
Hisp: Hispanic
M: male
MOR: mu-opioid receptor
iSUD: individuals with substance use disorders
Non-Hisp: Non-hispanic
NS: not significant
OUD: Opioid use disorder
OR: Odds ratios
Ref.: reference category
SDOH: social determinants of health
SPD: Schizophrenia or other psychotic disorders
SUD: substance use disorders

