## Supplementary material for "Psychiatric comorbidities in substance use disorders: Sex-based differences in a national real-world clinic sample": Results Supplement

**Table S1.** Demographics in the analyzed sample (individuals with primary diagnoses of opioid, alcohol or cannabis dependence)

| **Variable** | **Group** | **Total**  **N** | **Opioid Dependence** | | **Alcohol Dependence** | | **Cannabis Dependence** | | **Univariate Chi^2^ analysis**  **(df); p-value** |
| --- | --- | --- | --- | --- | --- | --- | --- | --- | --- |
| **Total** | | **58050** | **N** | **%** | **N** | **%** | **N** | **%** | **N/A** |
|  |  |  | **28808** | **49.6%** | **23281** | **40.1%** | **5961** | **10.3** |  |
| **Sex** | **Male** | **34244** | **15664** | **54.37%** | **14831** | **63.70%** | **3749** | **62.89%** | **504.9 (2); p<0.0001** |
|  | **Female** | **23806** | **13144** | **45.63%** | **8451** | **36.30%** | **2212** | **37.11%** |  |
| **Race** | **African-American / Black (AA)** | **6525** | **1711** | **5.94%** | **3079** | **13.23%** | **1735** | **29.11%** | **7000 (4); p<0.0001** |
|  | **American Indian /**  **Native Alaskan (AI)** | **6085** | **1162** | **4.03%** | **4701** | **20.19%** | **222** | **3.72%** |  |
|  | **White** | **45440** | **25935** | **90.03%** | **15501** | **66.58%** | **4004** | **67.17%** |  |
| **Ethnicity** | **Hispanic/Latino** | **28050** | **17989** | **62.44%** | **8947** | **38.43%** | **1114** | **18.69%** | **5309 (2); p<0.0001** |
|  | **Non-**  **Hispanic/Latino** | **30000** | **10819** | **37.56%** | **14334** | **61.57%** | **4847** | **81.31%** |  |
| **Age Bin** | **18-24** | **3821** | **1707** | **5.93%** | **1035** | **4.45%** | **1079** | **18.10%** | **5298 (8); p<0.0001** |
|  | **25-34** | **17788** | **10617** | **36.85%** | **5207** | **22.37%** | **1964** | **32.95%** |  |
|  | **35-44** | **16582** | **9291** | **32.25%** | **5779** | **24.82%** | **1512** | **25.36%** |  |
|  | **45-54** | **9533** | **4094** | **14.21%** | **4658** | **20.01%** | **781** | **13.10%** |  |
|  | **≥55** | **10326** | **3099** | **10.76%** | **6602** | **28.36%** | **625** | **10.48%** |  |

**Table S2:** Comorbidity outcomes in individuals with a primary diagnosis of opioid dependence

|  | | “n” | **Outcome: comorbidity*** | | | | | | | | | | | |
| --- | --- | --- | --- | --- | --- | --- | --- | --- | --- | --- | --- | --- | --- | --- |
|  |  |  | Anxiety | | Bipolar | | Depression | | Schizophrenia  or other psychotic disorders (SPD) | | Trauma / Stressor | | No comorbidity | |
| **Var.** | **Cond.** |  | N | % | N | % | N | % | N | % | N | % | N | % |
| Total | | 28808 | 4243 | 14.73 | 1353 | 4.70 | 3894 | 13.52 | 568 | 1.97 | 1979 | 6.87 | 16771 | 58.22 |
| Sex  Χ^2^ [df=5]  209.2  p<0.0001 | Male | 15664 | 2246 | 14.34 | 573 | 3.66 | 1987 | 12.69 | 380 | 2.43 | 949 | 6.06 | 9529 | 60.83 |
|  | Female | 13144 | 1997 | 15.19 | 780 | 5.93 | 1907 | 14.51 | 188 | 1.43 | 1030 | 7.84 | 7242 | 55.10 |
| Race  Sex  Χ^2^ [df=10]  73.05  p<0.0001 | AA | 1711 | 212 | 12.39 | 94 | 5.49 | 311 | 18.18 | 57 | 3.33 | 105 | 6.14 | 932 | 54.47 |
|  | AI | 1162 | 170 | 14.63 | 33 | 2.84 | 142 | 12.22 | 25 | 2.15 | 78 | 6.71 | 714 | 61.45 |
|  | White | 25935 | 3861 | 14.89 | 1226 | 4.73 | 3441 | 13.27 | 486 | 1.87 | 1796 | 6.93 | 15125 | 58.32 |
| Ethnicity  Sex  Χ^2^ [df=5]  455.9  p<0.000 | Hispanic/  Latino | 17989 | 3262 | 18.13 | 771 | 4.29 | 2346 | 13.04 | 309 | 1.72 | 1204 | 6.69 | 10097 | 56.13 |
|  | Not Hispanic/  Latino | 10819 | 981 | 9.07 | 582 | 5.38 | 1548 | 14.31 | 259 | 2.39 | 775 | 7.16 | 6674 | 61.69 |
| Age | 18-24 | 1707 | 343 | 20.09 | 88 | 5.16 | 293 | 17.16 | 29 | 1.70 | 115 | 6.74 | 839 | 49.15 |
|  | 25-34 | 10617 | 1709 | 16.10 | 522 | 4.92 | 1498 | 14.11 | 174 | 1.64 | 766 | 7.21 | 5948 | 56.02 |
|  | 35-44 | 9291 | 1407 | 15.14 | 400 | 4.31 | 1126 | 12.12 | 189 | 2.03 | 705 | 7.59 | 5464 | 58.81 |
|  | 45-54 | 4094 | 481 | 11.75 | 196 | 4.79 | 537 | 13.12 | 104 | 2.54 | 251 | 6.13 | 2525 | 61.68 |
|  | ≥55 | 3099 | 303 | 9.78 | 147 | 4.74 | 440 | 14.20 | 72 | 2.32 | 142 | 4.58 | 1995 | 64.38 |

**Note:** Individuals with all other comorbidities combined (not analyzed) were n=2062

**Table S3:** Psychiatric comorbidity in individuals with a primary diagnosis of alcohol dependence

|  | | “n” | **Outcome: comorbidity*** | | | | | | | | | | | |
| --- | --- | --- | --- | --- | --- | --- | --- | --- | --- | --- | --- | --- | --- | --- |
|  |  |  | Anxiety | | Bipolar | | Depression | | Schizophrenia  or other psychotic disorders (SPD) | | Trauma / Stressor | | No comorbidity | |
| **Var.** | **Cond.** |  | N | % | N | % | N | % | N | % | N | % | N | % |
| Total | | 23281 | 2790 | 11.98 | 1691 | 7.26 | 5069 | 21.77 | 1246 | 5.35 | 1718 | 7.38 | 10767 | 46.25 |
| Sex  Χ^2^ [df=5]  404.8  p<0.0001 | Male | 14831 | 1641 | 11.06 | 868 | 5.85 | 3007 | 20.28 | 901 | 6.08 | 965 | 6.51 | 7449 | 50.23 |
|  | Female | 8450 | 1149 | 13.60 | 823 | 9.74 | 2062 | 24.40 | 345 | 4.08 | 753 | 8.91 | 3318 | 39.27 |
| Race  Χ^2^ [df=10]  761.8  p<0.0001 | AA | 3079 | 192 | 6.24 | 217 | 7.05 | 592 | 19.23 | 321 | 10.43 | 210 | 6.82 | 1547 | 50.24 |
|  | AI | 4701 | 776 | 16.51 | 91 | 1.94 | 926 | 19.70 | 87 | 1.85 | 415 | 8.83 | 2406 | 51.18 |
|  | White | 15501 | 1822 | 11.75 | 1383 | 8.92 | 3551 | 22.91 | 838 | 5.41 | 1093 | 7.05 | 6814 | 43.96 |
| Ethnicity  Χ^2^ [df=5]  1544  p<0.0001 | Hispanic/  Latino | 8947 | 1862 | 20.81 | 510 | 5.70 | 2117 | 23.66 | 310 | 3.46 | 888 | 9.93 | 3260 | 36.44 |
|  | Not Hispanic/  Latino | 14334 | 928 | 6.47 | 1181 | 8.24 | 2952 | 20.59 | 936 | 6.53 | 830 | 5.79 | 7507 | 52.37 |
| Age | 18-24 | 1035 | 139 | 13.43 | 58 | 5.60 | 207 | 20.00 | 35 | 3.38 | 111 | 10.72 | 485 | 46.86 |
|  | 25-34 | 5207 | 812 | 15.59 | 327 | 6.28 | 1134 | 21.78 | 226 | 4.34 | 453 | 8.70 | 2255 | 43.31 |
|  | 35-44 | 5779 | 797 | 13.79 | 428 | 7.41 | 1213 | 20.99 | 303 | 5.24 | 483 | 8.36 | 2555 | 44.21 |
|  | 45-54 | 4658 | 566 | 12.15 | 399 | 8.57 | 1060 | 22.76 | 281 | 6.03 | 327 | 7.02 | 2025 | 43.47 |
|  | ≥55 | 6602 | 476 | 7.21 | 479 | 7.26 | 1455 | 22.04 | 401 | 6.07 | 344 | 5.21 | 3447 | 52.21 |

**Note:** Individuals with all other comorbidities combined (not analyzed) were n=2539

**Table S4:** Psychiatric comorbidity in individuals a primary diagnosis of cannabis dependence **(total n=5961)**

|  | | “n” | **Outcome: comorbidity*** | | | | | | | | | | | |
| --- | --- | --- | --- | --- | --- | --- | --- | --- | --- | --- | --- | --- | --- | --- |
|  |  |  | Anxiety | | Bipolar | | Depression | | Schizophrenia  or other psychotic disorders (SPD) | | Trauma / Stressor | | No comorbidity | |
| Var. | Cond. |  | N | % | N | % | N | % | N | % | N | % | N | % |
| Total | | 5961 | 518 | 8.69 | 737 | 12.36 | 1071 | 17.97 | 798 | 13.39 | 504 | 8.45 | 2333 | 39.14 |
| Sex  Χ^2^ [df=5]  316.5  p<0.0001 | Male | 3749 | 270 | 7.20 | 345 | 9.20 | 571 | 15.23 | 610 | 16.27 | 270 | 7.20 | 1683 | 44.89 |
|  | Female | 2212 | 248 | 11.21 | 392 | 17.72 | 500 | 22.60 | 188 | 8.50 | 234 | 10.58 | 650 | 29.39 |
| Race  Χ^2^ [df=10]  97.1  p<0.0001 | AA | 1735 | 97 | 5.59 | 166 | 9.57 | 280 | 16.14 | 296 | 17.06 | 144 | 8.30 | 752 | 43.34 |
|  | AI | 222 | 29 | 13.06 | 18 | 8.11 | 40 | 18.02 | 17 | 7.66 | 24 | 10.81 | 94 | 42.34 |
|  | White | 4004 | 392 | 9.79 | 553 | 13.81 | 751 | 18.76 | 485 | 12.11 | 336 | 8.39 | 1487 | 37.14 |
| Ethnicity  Χ^2^ [df=5]  138.2  p<0.0001 | Hispanic/  Latino | 1114 | 173 | 15.53 | 118 | 10.59 | 210 | 18.85 | 148 | 13.29 | 139 | 12.48 | 326 | 29.26 |
|  | Not Hispanic/  Latino | 4847 | 345 | 7.12 | 619 | 12.77 | 861 | 17.76 | 650 | 13.41 | 365 | 7.53 | 2007 | 41.41 |
| Age | 18-24 | 1079 | 126 | 11.68 | 129 | 11.96 | 243 | 22.52 | 97 | 8.99 | 99 | 9.18 | 385 | 35.68 |
|  | 25-34 | 1964 | 199 | 10.13 | 267 | 13.59 | 358 | 18.23 | 269 | 13.70 | 172 | 8.76 | 699 | 35.59 |
|  | 35-44 | 1512 | 113 | 7.47 | 166 | 10.98 | 236 | 15.61 | 195 | 12.90 | 116 | 7.67 | 686 | 45.37 |
|  | 45-54 | 781 | 45 | 5.76 | 99 | 12.68 | 112 | 14.34 | 116 | 14.85 | 72 | 9.22 | 337 | 43.15 |
|  | ≥55 | 625 | 35 | 5.60 | 76 | 12.16 | 122 | 19.52 | 121 | 19.36 | 45 | 7.20 | 226 | 36.16 |

**Note:** Individuals with all other comorbidities combined (not analyzed) were n=649

**Figure S1:** Relative frequency of different comorbidity outcomes in all persons (males and females combined) with opioid, alcohol or cannabis dependence as their primary diagnoses. Data are further stratified by sex, race and ethnicity (Fig. 1), and tabulated in Tables S2-4). These data were analyzed in three separate multinomial logistic regressions (Figs. 2-4, and Tables S5-7).

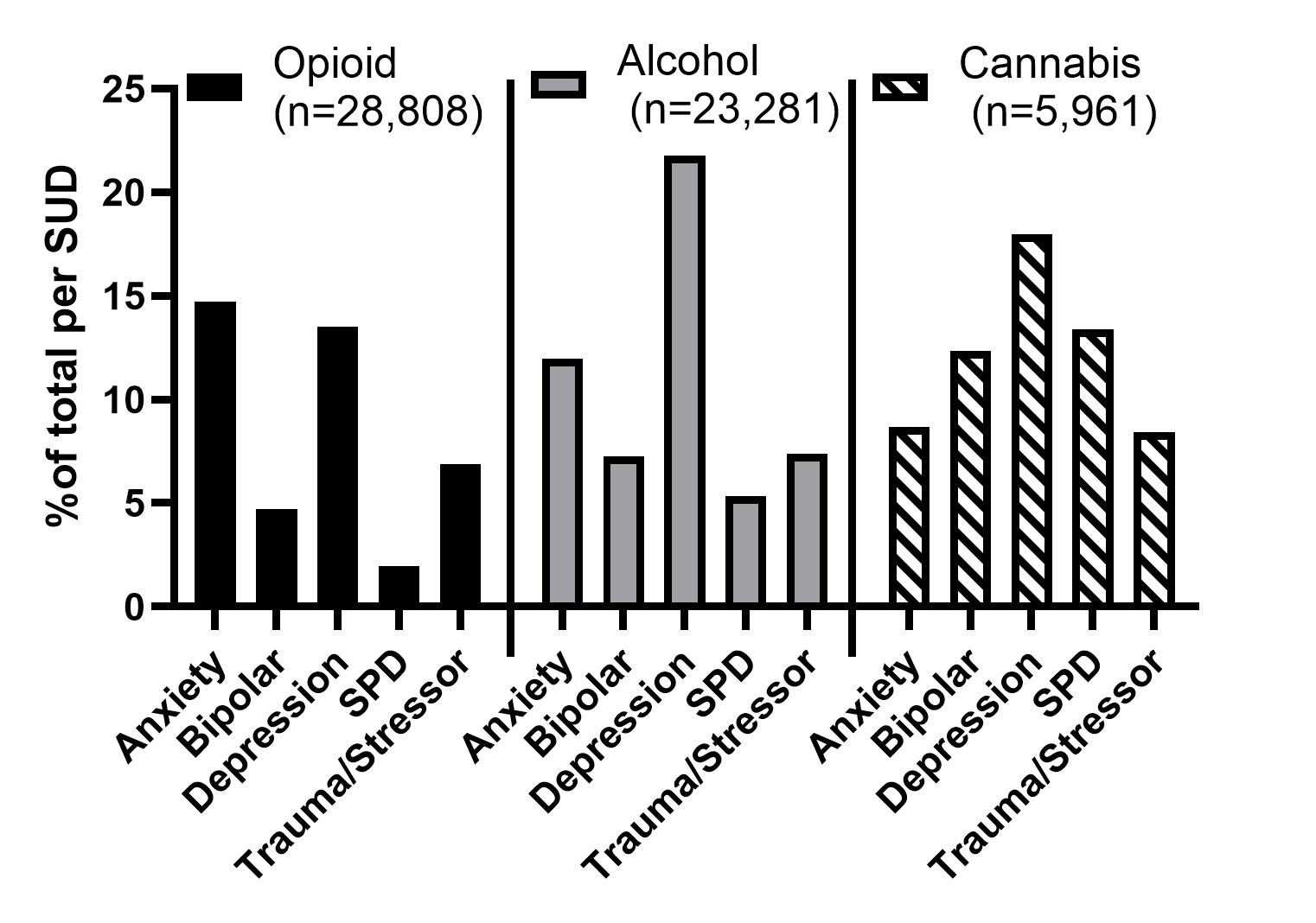

**Table S5.** Multinomial logistic regression for comorbidity outcomes in individuals with a primary diagnosis of opioid dependence (data in Table S2).

Likelihood ratio tests are provided in the footnote*

| Outcome | Variable | Condition | Regression Parameters | | | p-value | Odds ratio | | |
| --- | --- | --- | --- | --- | --- | --- | --- | --- | --- |
|  |  | | Beta | Lower 95% CI | Upper 95% CI |  | Odds ratio (OR) | Lower 95% CI | Upper 95% CI |
| Anxiety | Intercept | | -1.60 | -1.68 | -1.52 |  |  |  |  |
|  | Sex | Male | -0.08 | -0.12 | -0.05 | 0.000003 | 0.92 | 0.89 | 0.95 |
|  | Race | AA | 0.02 | -0.10 | 0.13 | 0.778 | 1.02 | 0.91 | 1.14 |
|  |  | AI | -0.07 | -0.20 | 0.05 | 0.257 | 0.93 | 0.82 | 1.05 |
|  | Ethnicity | Hispanic | 0.38 | 0.35 | 0.42 | <0.000001 | 1.47 | 1.41 | 1.53 |
|  | Age | 18-24 | 0.47 | 0.36 | 0.58 | <0.000001 | 1.60 | 1.44 | 1.78 |
|  |  | 25-34 | 0.15 | 0.09 | 0.21 | 0.000001 | 1.16 | 1.09 | 1.23 |
|  |  | 35-44 | 0.07 | 0.01 | 0.14 | 0.0207 | 1.08 | 1.01 | 1.15 |
|  |  | 45-54 | -0.24 | -0.32 | -0.15 | <0.000001 | 0.79 | 0.72 | 0.86 |
| Bipolar | Intercept | | -2.62 | -2.76 | -2.47 |  |  |  |  |
|  | Sex | Male | -0.30 | -0.35 | -0.24 | <0.000001 | 0.74 | 0.70 | 0.79 |
|  | Race | AA | 0.35 | 0.16 | 0.53 | 0.0002 | 1.42 | 1.18 | 1.70 |
|  |  | AI | -0.49 | -0.74 | -0.25 | 0.0001 | 0.61 | 0.48 | 0.78 |
|  | Ethnicity | Hispanic | -0.07 | -0.13 | -0.01 | 0.0168 | 0.93 | 0.88 | 0.99 |
|  | Age | 18-24 | 0.26 | 0.07 | 0.44 | 0.0063 | 1.29 | 1.07 | 1.55 |
|  |  | 25-34 | 0.06 | -0.03 | 0.16 | 0.192 | 1.07 | 0.97 | 1.18 |
|  |  | 35-44 | -0.13 | -0.23 | -0.02 | 0.0175 | 0.88 | 0.79 | 0.98 |
|  |  | 45-54 | -0.07 | -0.20 | 0.06 | 0.287 | 0.93 | 0.82 | 1.06 |
| Depression | Intercept | | -1.36 | -1.44 | -1.29 |  |  |  |  |
|  | Sex | Male | -0.12 | -0.16 | -0.09 | <0.000001 | 0.89 | 0.86 | 0.92 |
|  | Race | AA | 0.30 | 0.20 | 0.41 | <0.000001 | 1.36 | 1.22 | 1.51 |
|  |  | AI | -0.25 | -0.37 | -0.12 | 0.0002 | 0.78 | 0.69 | 0.89 |
|  | Ethnicity | Hispanic | 0.00 | -0.04 | 0.03 | 0.942 | 1.00 | 0.96 | 1.04 |
|  | Age | 18-24 | 0.36 | 0.25 | 0.47 | <0.000001 | 1.43 | 1.28 | 1.60 |
|  |  | 25-34 | 0.04 | -0.02 | 0.10 | 0.238 | 1.04 | 0.98 | 1.10 |
|  |  | 35-44 | -0.16 | -0.22 | -0.09 | 0.000002 | 0.85 | 0.80 | 0.91 |
|  |  | 45-54 | -0.13 | -0.22 | -0.05 | 0.0020 | 0.88 | 0.81 | 0.95 |
| SPD | Intercept | | -3.20 | -3.37 | -3.02 |  |  |  |  |
|  | Sex | Male | 0.21 | 0.12 | 0.30 | 0.000003 | 1.24 | 1.13 | 1.35 |
|  | Race | AA | 0.36 | 0.14 | 0.59 | 0.0017 | 1.44 | 1.15 | 1.80 |
|  |  | AI | -0.11 | -0.40 | 0.17 | 0.435 | 0.89 | 0.67 | 1.19 |
|  | Ethnicity | Hispanic | -0.10 | -0.19 | -0.02 | 0.0180 | 0.90 | 0.83 | 0.98 |
|  | Age | 18-24 | -0.02 | -0.33 | 0.29 | 0.90 | 0.98 | 0.72 | 1.33 |
|  |  | 25-34 | -0.17 | -0.33 | -0.02 | 0.0314 | 0.84 | 0.72 | 0.98 |
|  |  | 35-44 | 0.01 | -0.15 | 0.16 | 0.944 | 1.01 | 0.86 | 1.17 |
|  |  | 45-54 | 0.18 | -0.01 | 0.36 | 0.0624 | 1.19 | 0.99 | 1.43 |
| Trauma /  Stressor | Intercept | | -2.24 | -2.36 | -2.13 |  |  |  |  |
|  | Sex | Male | -0.18 | -0.23 | -0.13 | <0.000001 | 0.84 | 0.80 | 0.88 |
|  | Race | AA | 0.03 | -0.13 | 0.19 | 0.7058 | 1.03 | 0.88 | 1.21 |
|  |  | AI | -0.08 | -0.25 | 0.09 | 0.350 | 0.92 | 0.78 | 1.09 |
|  | Ethnicity | Hispanic | 0.01 | -0.04 | 0.06 | 0.685 | 1.01 | 0.96 | 1.06 |
|  | Age | 18-24 | 0.22 | 0.06 | 0.39 | 0.0077 | 1.25 | 1.06 | 1.47 |
|  |  | 25-34 | 0.16 | 0.07 | 0.24 | 0.0003 | 1.17 | 1.08 | 1.28 |
|  |  | 35-44 | 0.16 | 0.07 | 0.24 | 0.0004 | 1.17 | 1.07 | 1.28 |
|  |  | 45-54 | -0.11 | -0.22 | 0.01 | 0.0799 | 0.90 | 0.80 | 1.01 |

***Likelihood ratio test (type 3), examining unique contributions of each independent variable:** Age [DF=20], log-likelihood= -36281.5, Chi-square=261.5; ethnicity [DF=5], log-likelihood= -36377.8, Chi-square=454.1; race [DF=10] log-likelihood= -36182.9, Chi-square=64.3; sex [DF=5]	log-likelihood= -36257.5, Chi-square=213.6. Each variable had p<0.001.

**Table S6.** Multinomial logistic regression for comorbidity outcomes in individuals with a primary diagnosis of alcohol dependence (data in Table S3). Likelihood ratio tests are provided in the footnote**

| Outcome | Variable | Condition | Regression Parameters | | | p-value | Odds ratio | | |
| --- | --- | --- | --- | --- | --- | --- | --- | --- | --- |
|  |  | | Beta | Lower 95% CI | Upper 95% CI |  | Odds ratio (OR) | Lower 95% CI | Upper 95% CI |
| Anxiety | Intercept | | -1.42 | -1.49 | -1.35 |  |  |  |  |
|  | Sex | Male | -0.21 | -0.25 | -0.16 | <0.00001 | 0.81 | 0.78 | 0.85 |
|  | Race | AA | -0.18 | -0.29 | -0.07 | 0.001440 | 0.84 | 0.75 | 0.93 |
|  |  | AI | -0.09 | -0.17 | -0.01 | 0.030 | 0.91 | 0.84 | 0.99 |
|  | Ethnicity | Hispanic | 0.74 | 0.69 | 0.78 | <0.00001 | 2.09 | 1.99 | 2.19 |
|  | Age | 18-24 | -0.11 | -0.27 | 0.05 | 0.160 | 0.89 | 0.76 | 1.05 |
|  |  | 25-34 | 0.22 | 0.14 | 0.30 | <0.00001 | 1.24 | 1.15 | 1.35 |
|  |  | 35-44 | 0.16 | 0.07 | 0.24 | 0.000246 | 1.17 | 1.07 | 1.27 |
|  |  | 45-54 | 0.11 | 0.02 | 0.20 | 0.017 | 1.12 | 1.02 | 1.23 |
| Bipolar | Intercept | | -2.19 | -2.29 | -2.09 | <0.00001 |  |  |  |
|  | Sex | Male | -0.38 | -0.43 | -0.33 | <0.00001 | 0.68 | 0.65 | 0.72 |
|  | Race | AA | 0.40 | 0.27 | 0.52 | <0.00001 | 1.48 | 1.31 | 1.68 |
|  |  | AI | -1.10 | -1.26 | -0.95 | <0.00001 | 0.33 | 0.29 | 0.39 |
|  | Ethnicity | Hispanic | 0.10 | 0.04 | 0.16 | 0.000606 | 1.11 | 1.04 | 1.18 |
|  | Age | 18-24 | -0.19 | -0.42 | 0.03 | 0.092 | 0.82 | 0.66 | 1.03 |
|  |  | 25-34 | 0.04 | -0.07 | 0.16 | 0.453 | 1.04 | 0.93 | 1.17 |
|  |  | 35-44 | 0.14 | 0.03 | 0.25 | 0.010 | 1.15 | 1.03 | 1.28 |
|  |  | 45-54 | 0.21 | 0.10 | 0.32 | 0.000136 | 1.24 | 1.11 | 1.38 |
| Depression | Intercept | | -0.74 | -0.79 | -0.69 | <0.00001 |  |  |  |
|  | Sex | Male | -0.21 | -0.25 | -0.18 | <0.00001 | 0.81 | 0.78 | 0.84 |
|  | Race | AA | 0.04 | -0.04 | 0.11 | 0.323 | 1.04 | 0.96 | 1.11 |
|  |  | AI | -0.27 | -0.34 | -0.21 | <0.00001 | 0.76 | 0.71 | 0.81 |
|  | Ethnicity | Hispanic | 0.29 | 0.25 | 0.33 | <0.00001 | 1.34 | 1.29 | 1.39 |
|  | Age | 18-24 | -0.15 | -0.29 | -0.02 | 0.026 | 0.86 | 0.75 | 0.98 |
|  |  | 25-34 | 0.06 | -0.01 | 0.13 | 0.082 | 1.07 | 0.99 | 1.14 |
|  |  | 35-44 | 0.03 | -0.04 | 0.09 | 0.482 | 1.03 | 0.96 | 1.10 |
|  |  | 45-54 | 0.12 | 0.05 | 0.19 | 0.001500 | 1.13 | 1.05 | 1.21 |
| SPD | Intercept | | -2.40 | -2.50 | -2.29 | <0.00001 |  |  |  |
|  | Sex | Male | 0.06 | 0.00 | 0.13 | 0.057 | 1.07 | 1.00 | 1.14 |
|  | Race | AA | 0.77 | 0.66 | 0.88 | <0.00001 | 2.16 | 1.93 | 2.42 |
|  |  | AI | -1.01 | -1.17 | -0.86 | <0.00001 | 0.36 | 0.31 | 0.42 |
|  | Ethnicity | Hispanic | 0.02 | -0.05 | 0.09 | 0.622 | 1.02 | 0.95 | 1.09 |
|  | Age | 18-24 | -0.28 | -0.56 | 0.01 | 0.055 | 0.76 | 0.57 | 1.01 |
|  |  | 25-34 | 0.04 | -0.10 | 0.18 | 0.583 | 1.04 | 0.91 | 1.19 |
|  |  | 35-44 | 0.13 | 0.01 | 0.26 | 0.040 | 1.14 | 1.01 | 1.29 |
|  |  | 45-54 | 0.19 | 0.06 | 0.32 | 0.004 | 1.21 | 1.06 | 1.38 |
| Trauma /  Stressor | Intercept | | -1.68 | -1.75 | -1.61 | <0.00001 |  |  |  |
|  | Sex | Male | -0.26 | -0.32 | -0.21 | <0.00001 | 0.77 | 0.73 | 0.81 |
|  | Race | AA | 0.12 | 0.02 | 0.23 | 0.026 | 1.13 | 1.02 | 1.26 |
|  |  | AI | -0.23 | -0.32 | -0.13 | 0.000003 | 0.80 | 0.73 | 0.88 |
|  | Ethnicity | Hispanic | 0.44 | 0.38 | 0.50 | <0.00001 | 1.55 | 1.47 | 1.64 |
|  | Age | 18-24 | 0.18 | 0.00 | 0.35 | 0.044 | 1.19 | 1.00 | 1.42 |
|  |  | 25-34 | 0.12 | 0.02 | 0.22 | 0.016 | 1.13 | 1.02 | 1.25 |
|  |  | 35-44 | 0.11 | 0.01 | 0.21 | 0.025 | 1.12 | 1.01 | 1.23 |
|  |  | 45-54 | -0.02 | -0.13 | 0.09 | 0.731 | 0.98 | 0.88 | 1.09 |

****Likelihood ratio test (type 3), examining unique contributions of each independent variable:** Age [DF=20], log-likelihood= -33229.4, Chi-square=178.4; ethnicity [DF=5], log-likelihood= -33721.8, Chi-square=1163.2; race [DF=10] log-likelihood= -33470.1, Chi-square=659.8; sex [DF=5]	log-likelihood= -33323.4, Chi-square=366.4. Each predictor had p<0.01.

**Table S7.** Multinomial logistic regression for comorbidity outcomes in individuals with a primary diagnosis of cannabis dependence (data in Table S4). Likelihood ratio tests are provided in the footnote***

| Outcome | Variable | Condition | Regression Parameters | | | p-value | Odds ratio | | |
| --- | --- | --- | --- | --- | --- | --- | --- | --- | --- |
|  |  | | Beta | Lower 95% CI | Upper 95% CI |  | Odds ratio (OR) | Lower 95% CI | Upper 95% CI |
| Anxiety | Intercept | | -1.38 | -1.56 | -1.20 |  |  |  |  |
|  | Sex | Male | -0.40 | -0.50 | -0.30 | <0.000001 | 0.67 | 0.61 | 0.74 |
|  | Race | AA | -0.25 | -0.46 | -0.04 | 0.019 | 0.78 | 0.63 | 0.96 |
|  |  | AI | -0.05 | -0.36 | 0.25 | 0.742 | 0.95 | 0.70 | 1.28 |
|  | Ethnicity | Hispanic | 0.50 | 0.38 | 0.61 | <0.000001 | 1.65 | 1.46 | 1.84 |
|  | Age | 18-24 | 0.40 | 0.20 | 0.60 | 0.00007 | 1.49 | 1.22 | 1.82 |
|  |  | 25-34 | 0.32 | 0.15 | 0.49 | 0.00019 | 1.38 | 1.16 | 1.63 |
|  |  | 35-44 | -0.13 | -0.33 | 0.06 | 0.180 | 0.88 | 0.72 | 1.06 |
|  |  | 45-54 | -0.35 | -0.62 | -0.08 | 0.012 | 0.70 | 0.54 | 0.92 |
| Bipolar | Intercept | | -1.27 | -1.47 | -1.08 |  |  |  |  |
|  | Sex | Male | -0.52 | -0.61 | -0.44 | <0.000001 | 0.59 | 0.54 | 0.64 |
|  | Race | AA | -0.03 | -0.24 | 0.18 | 0.794 | 0.97 | 0.79 | 1.20 |
|  |  | AI | -0.37 | -0.72 | -0.02 | 0.041 | 0.69 | 0.49 | 0.98 |
|  | Ethnicity | Hispanic | 0.06 | -0.06 | 0.17 | 0.364 | 1.06 | 0.94 | 1.19 |
|  | Age | 18-24 | 0.07 | -0.12 | 0.25 | 0.475 | 1.07 | 0.89 | 1.28 |
|  |  | 25-34 | 0.18 | 0.04 | 0.33 | 0.012 | 1.20 | 1.04 | 1.39 |
|  |  | 35-44 | -0.24 | -0.40 | -0.07 | 0.004 | 0.79 | 0.67 | 0.93 |
|  |  | 45-54 | -0.04 | -0.24 | 0.16 | 0.670 | 0.96 | 0.79 | 1.17 |
| Depression | Intercept | | -0.69 | -0.83 | -0.54 |  |  |  |  |
|  | Sex | Male | -0.40 | -0.47 | -0.32 | <0.000001 | 0.67 | 0.63 | 0.73 |
|  | Race | AA | -0.01 | -0.17 | 0.15 | 0.906 | 0.99 | 0.84 | 1.16 |
|  |  | AI | -0.18 | -0.44 | 0.08 | 0.179 | 0.84 | 0.64 | 1.08 |
|  | Ethnicity | Hispanic | 0.17 | 0.07 | 0.27 | 0.001 | 1.19 | 1.07 | 1.31 |
|  | Age | 18-24 | 0.31 | 0.16 | 0.46 | 0.00007 | 1.36 | 1.17 | 1.58 |
|  |  | 25-34 | 0.10 | -0.03 | 0.23 | 0.134 | 1.11 | 0.97 | 1.26 |
|  |  | 35-44 | -0.26 | -0.40 | -0.12 | 0.00032 | 0.77 | 0.67 | 0.89 |
|  |  | 45-54 | -0.29 | -0.48 | -0.11 | 0.002 | 0.75 | 0.62 | 0.90 |
| Outcome | Variable | Condition |  |  |  |  |  |  |  |
| SPD | Intercept | | -1.18 | -1.37 | -0.98 |  |  |  |  |
|  | Sex | Male | 0.11 | 0.02 | 0.21 | 0.017 | 1.12 | 1.02 | 1.23 |
|  | Race | AA | 0.39 | 0.19 | 0.60 | 0.00013 | 1.48 | 1.21 | 1.82 |
|  |  | AI | -0.57 | -0.92 | -0.21 | 0.002 | 0.57 | 0.40 | 0.81 |
|  | Ethnicity | Hispanic | 0.24 | 0.13 | 0.35 | 0.00003 | 1.27 | 1.14 | 1.42 |
|  | Age | 18-24 | -0.34 | -0.53 | -0.15 | 0.001 | 0.71 | 0.59 | 0.86 |
|  |  | 25-34 | 0.09 | -0.05 | 0.23 | 0.199 | 1.09 | 0.95 | 1.26 |
|  |  | 35-44 | -0.21 | -0.36 | -0.05 | 0.008 | 0.81 | 0.70 | 0.95 |
|  |  | 45-54 | -0.01 | -0.19 | 0.18 | 0.941 | 0.99 | 0.83 | 1.20 |
| Trauma /  Stressor | Intercept | | -1.27 | -1.45 | -1.09 |  |  |  |  |
|  | Sex | Male | -0.39 | -0.49 | -0.29 | <0.000001 | 0.68 | 0.61 | 0.75 |
|  | Race | AA | 0.07 | -0.14 | 0.27 | 0.515 | 1.07 | 0.87 | 1.31 |
|  |  | AI | -0.17 | -0.49 | 0.15 | 0.301 | 0.84 | 0.61 | 1.16 |
|  | Ethnicity | Hispanic | 0.41 | 0.29 | 0.53 | <0.00001 | 1.51 | 1.34 | 1.70 |
|  | Age | 18-24 | 0.12 | -0.08 | 0.33 | 0.239 | 1.13 | 0.92 | 1.39 |
|  |  | 25-34 | 0.11 | -0.06 | 0.27 | 0.223 | 1.12 | 0.94 | 1.31 |
|  |  | 35-44 | -0.20 | -0.39 | -0.01 | 0.035 | 0.82 | 0.68 | 0.99 |
|  |  | 45-54 | 0.03 | -0.19 | 0.26 | 0.770 | 1.03 | 0.83 | 1.30 |

*****Likelihood ratio test (type 3), examining unique contributions of each independent variable:** Age [DF=20], log-likelihood= -9428.8, Chi-square=108.6; ethnicity [DF=5], log-likelihood= -9424.4, Chi-square=99.8; race [DF=10] log-likelihood= -9408.9, Chi-square=69.9; sex [DF=5]	log-likelihood= -9519.3, Chi-square=289.6. Each predictor had p<0.001.
