## Supplementary material for "Psychiatric comorbidities in substance use disorders: Sex-based differences in a national real-world clinic sample": Participant filtering table

**Participant Selection Table:** Data from Mental Health-Client Level Data for 2022; Substance Abuse and Mental Health Services Administration (SAMHSA).

Raw data, sample sizes, and variable codebook available at:

https://www.samhsa.gov/data/data-we-collect/mh-cld/datafiles

| **Participant selection stage** | **“N”** | **“N” remaining** | **Missing** | **Number excluded** | **Notes** |
| --- | --- | --- | --- | --- | --- |
| Whole sample | 6,957,919 | 6,957,919 | N/A | N/A | N/A |
| Participants with primary mental health diagnosis (variable MH1) of  “Alcohol or substance use disorders” | | 236,450 | 1,058,221 | N/A | N/A |
| Participants with current substance use diagnosis (variable SUB) of either: “opioid dependence”, “alcohol dependence” or “cannabis dependence” | | 92,571 | 38,665 | 105,214 | Other SUD diagnoses excluded |
| Participants with specific psychiatric comorbidity as their secondary mental health diagnosis (variable MH2), or no comorbidity. The selected comorbidity groups were: “anxiety disorders”, “bipolar disorders”, “depressive disorders”, “schizophrenia or other psychotic disorders”, “trauma and stressor-related disorders”. No comorbidity status was based on number of diagnoses=1 (variable NUMMHS) | | 85,071 | N/A | 7,500 | Comorbidities other than anxiety, bipolar, depression, SPD or trauma.stressor disorders excluded |
| Participants aged: ≥18 (variable AGE);  Recoded as age bins: 18-24; 25-34; 35-44; 45-54 and ≥55 | | 83,614 | 1 | 1,456 | Ages ≤18 excluded |
| Participants of latin/hispanic ethnicity (variable ETHNIC); recoded into one category), versus participants not of latin/hispanic ethnicity | | 66,896 | 16,718 | N/A | N/A |
| Participants who are in the following racial categories (variable RACE) black/african-american, American indian or Alaska native, or white | | 58,229 | 2,429 | 6,238 | Other racial groups, or those reporting more than one racial group excluded |
| Participants of male or female sex (variable GENDER) | | 58,052 | 177 | N/A | N/A |
| Participants in Midwest, Northeast, South and West U.S. Census Regions (variable REGION) | | 58,050 | N/A | 2 | Participants in “other jurisdictions” excluded. |
